# Cerebellar rTMS in PSP: a double blind sham-controlled study using mobile health technology

**DOI:** 10.1101/2020.11.09.20226068

**Authors:** Andrea Pilotto, Maria Cristina Rizzetti, Alberto Lombardi, Clint Hansen, Michele Biggi, Giacomo Verzeroli G, Antonella Martinelli, Robbin Romijnders, Barbara Borroni, Walter Maetzler, Alessandro Padovani

**Affiliations:** Neurology Unit, Department of Clinical and Experimental Sciences, University of Brescia, Brescia, Italy; Parkinson’s disease Rehabilitation unit, FERB Onlus Trescore Balneario, Bergamo, Italy; Physical Therapy Unit, Milano Bicocca University, Bergamo, Italy; Department of Neurology, Christian-Albrechts-University of Kiel, Kiel, Germany

**Keywords:** Progressive supranuclear palsy, repetitive transcranial magnetic stimulation, postural instability, mobile health technology

## Abstract

There are no effective treatments for postural instability and falls in Progressive Supranuclear Palsy (PSP). Objective of the study was to test the efficacy of theta burst repetitive transcranial magnetic stimulation (rTMS) on postural instability in PSP. Twenty probable PSP patients underwent a session of sham or cerebellar rTMS in a cross-over design. Before and after stimulation, static balance was evaluated with instrumented (lower back accelerometer, Rehagait®, Hasomed, Germany) 30-seconds-trials in semitandem and tandem positions. In tandem and semitandem tasks, active stimulation was associated with longer time without support falls (both p=0.04). In the same tasks, device-extracted parameters revealed significant improvement in area (p=0.007), velocity (p=0.005) and acceleration and jerkiness of sway (p=0.008) in real versus sham stimulation. Cerebellar rTMS thus showed a significant effect on stability in PSP patients, when assessed with mobile digital technology, in a double-blind design. These results should motivate larger and longer trials using non-invasive brain stimulation for PSP patients.

## INTRODUCTION

Progressive Supranuclear Palsy (PSP) is a neurodegenerative disorder characterised by akinetic rigid syndrome with ocular motor dysfunction, early postural instability and falls [1–3].

Despite potential limited benefit from dopaminergic drugs, there are still no effective treatments available for postural instability and falls. Recent imaging and neuropathology studies revealed a reduced volume of the cerebellum with Tau accumulation in PSP patients [4,5]. These evidences suggest that cerebellum may be a potential target for non-invasive stimulation, as already demonstrated for neurodegenerative ataxias [6].

Accordingly, neurophysiological studies demonstrated an impairment in functional connectivity between the cerebellar hemispheres and contralateral primary motor cortex (cerebellar brain-inhibition, CBI) [7–9]. A preliminary, open-label trial with 10 PSP patients showed an improvement of CBI using theta burst repetitive cerebellar transcranial magnetic stimulation (rTMS) [9]. Moreover, a case study showed improvement of posturography parameters secondary to cerebellar stimulation in two PSP patients [10].

Based on these promising results, we aimed at evaluating the effect of a single session cerebellar rTMS in PSP patients. We applied a double blind sham-controlled crossover design, including a standardised assessment of static balance using mobile health technology [11].

## METHODS

### Study cohort

PSP patients were recruited from the Parkinson’s disease Rehabilitation Unit at FERB ONLUS, Trescore Balneario, Bergamo, Italy. The diagnostic assessment included a review of the medical history, a neurological examination including the PSP Rating Scale (PSPRS) [12] and a comprehensive cognitive and behavioural assessment [13]. Inclusion criteria were as follows: 1) clinical diagnosis of probable PSP according to current clinical criteria [3], 2) the ability to stand alone without support, and 3) the ability to walk at least three meters without aid. Exclusion criteria were as follows: 1) dementia and 2) any contraindication to perform brain stimulation [8]. All subjects gave written informed consent prior to participation. The local ethics committee approved the study (protocol 193/16), recorded as NCT04222218 in clinicaltrial.gov. The study was performed in accordance with the Declaration of Helsinki.

### Repetitive transcranial Magnetic stimulation (rTMS)

Each patient received both rTMS and sham cerebellar single-session stimulation in randomized order in two different sessions, separated by at least two weeks. The patient and the examiner were blind to the type of rTMS delivered, which was applied by another experimenter. Repetitive cerebellar theta burst stimulation was performed by Duo-Mag XT100 (Deymed -Horonov, Czech republic) according to the protocol described by Brusa and coauthors [9]. The coil was placed tangentially to the skull over the lateral cerebellum 1 cm inferior and 3 cm right to the inion. Previous MRI studies showed that this stimulation targets the posterior and superior lobules of the lateral cerebellum [9,14]. Three 50-Hz pulses were repeated at a rate of 5 Hz; 20 trains of 10 bursts given with 8-s intervals for a total of 600 pulses and for total time of 240 seconds [9,15]. Intensity of rTMS was set at the 80% of the Resting Motor Threshold obtained in the left motor cortex for each subject. For sham simulation, a spacer was attached to the coil; the stimulation parameters, the coil position and the sound were identical to the active condition.

### Dynamic mobility and mobile health technology-instrumented static balance assessment

All subjects underwent a clinical evaluation including the Tinetti test [16], the Short Physical Performance Battery (SPPB) [17]), the Timed Up and Go test and the Functional Reach test (FR) [18] before and after stimulation. Static balance was tested before and after each stimulation with four tasks of 30s duration, respectively: tandem and semitandem stance with eyes closed and with eyes open (Supplementary Figure 1). As primary outcome measure, the time in seconds during the task without falling was recorded.

As secondary outcomes, an inertial sensor unit (IMU) with 100 Hz sampling frequency (Rehagait®, Hasomed GmbH, Magdeburg, Germany) was fixed at the level of the third lumbar spine segment close to the centre of mass [18]. Acceleration signals were processed and calculated as previously described [11,19,20]. The following sway parameters were extracted: area, mean velocity, mean acceleration (root mean square - RMS), jerk (indicating smoothness of compensatory movements [5]), and mean frequency [21]. Mean velocity, RMS and jerk were calculated for both anteroposterior (AP) and mediolateral (ML) directions.

### Statistical analyses

Differences in baseline performances between real and sham trials were evaluated using Mann-Whitney test. A two-way repeated-measures analysis of variance (ANOVA) was run to determine the effect of the different treatments over time on assessment, adjusted for baseline values and the sequence of stimulation (real-sham vs sham-real). Data are expressed as mean and standard deviation (SD), unless otherwise stated. SPSS software (version 21; SPSS, Inc., Chicago, IL) was used.

## RESULTS

### Study cohort and rTMS

Twenty PSP patients entered the study (mean age 74 ± 4 years, 13 males and 7 females). The mean disease duration was 3.8 ± 1.2 years, the mean score on the PSP rating scale was 29 ± 5 points, participants received dopaminergic medication with a mean levodopa equivalent dose of 417 ± 89 mg/day. All patients presented with postural instability, as reflected by specific PSPRS items and Tinetti scale. The rTMS protocol was well tolerated by all participants; side effects were neither reported nor observed during and after the stimulations.

There was no statistically significant association between type of stimulation and perception of patients (p=0.89, Fisher’s exact test), suggesting that real rTMS could not be distinguished from sham stimulation.

### Dynamic mobility and mobile health technology-instrumented static balance assessment

All patients were able to complete the 30s semitandem/tandem stance trials with eyes open, respectively. Sixteen participants completed the semitandem trial with eyes closed, and 14 the tandem trial with eyes closed. No differences in baseline performances in instrumented tests were detected for each task between real and sham stimulation. In both eyes closed conditions, the participants were able to stay longer without support after the real rTMS, compared to sham stimulation (time, Table 1). Moreover, in the tandem stance with eyes closed condition, the real intervention showed an improvement of the following parameters, compared to sham intervention: area, velocity, velocity in anterior-posterior direction, acceleration, and jerk in the medio-lateral direction. In the semitandem stance with eyes closed condition, the real trial showed an increase of velocity in the medio-lateral direction and a decrease of global and medio-lateral jerkiness, compared to sham intervention (Table 1).

**Table 1.**

| Variable | Pre-SHAM | Post-SHAM | Pre-REAL | Post-REAL | p |
| --- | --- | --- | --- | --- | --- |
| ***Tandem eyes closed (n=14)*** |  |  |  |  |  |
| 30s task completed, n | 9 | 7 | 8 | 7 | 0.8 |
| TIME, s | 21.1 ± 12.2 | 19.7 ± 13.7 | 20.9 ± 11.8 | 22.5 ± 11.2 | **0.046** |
| AREA, mm<sup>2</sup> | 6.55 ± 5.71 | 12.81 ± 11.66 | 25.42 ± 24.06 | 12.79 ± 15.26 | **0.007** |
| VELOCITY |  |  |  |  |  |
| MV, mm/s | 158.50 ± 33.37 | 206.89 ± 102.57 | 289.38 ± 242.22 | 202.87 ± 85.49 | **0.044** |
| MV-AP, mm/s | 20.04 ± 10.14 | 27.76 ± 18.67 | 35.97 ± 23.34 | 24.42 ± 13.72 | **0.009** |
| MV-ML, mm/s | 77.99 ± 65.36 | 103.62 ± 94.79 | 134.97 ± 199.56 | 89.87 ± 71.01 | 0.299 |
| ACCELERATION |  |  |  |  |  |
| ACC, mm/s<sup>2</sup> | 27.01 ± 12.16 | 36.45 ± 20.24 | 48.80 ± 29.57 | 34.90 ± 19.13 | **0.005** |
| ACC-AP, mm/s<sup>2</sup> | 62.10 ± 41.89 | 68.20 ± 48.91 | 84.01 ± 72.00 | 76.32 ± 46.67 | 0.544 |
| ACC-ML, mm/s<sup>2</sup> | 0.97 ± 0.80 | 1.05 ± 0.80 | 1.94 ± 2.84 | 2.98 ± 5.49 | 0.575 |
| JERK |  |  |  |  |  |
| JERK, mm/s<sup>3</sup> | 6.66 ± 6.60 | 8.14 ± 6.36 | 19.86 ± 30.36 | 7.42 ± 5.33 | 0.069 |
| JERK-AP, mm/s<sup>3</sup> | 1.02 ± 0.76 | 1.69 ± 2.06 | 1.99 ± 2.70 | 1.92 ± 2.97 | 0.514 |
| JERK-ML, mm/s<sup>3</sup> | 17.53 ± 8.25 | 22.92 ± 9.97 | 31.68 ± 20.66 | 24.20 ± 14.72 | **0.040** |
| FREQUENCY |  |  |  |  |  |
| MF, Hz | 1.34 ± 0.36 | 1.25 ± 0.37 | 1.29 ± 0.51 | 1.27 ± 0.34 | 0.647 |
| ***Semitandem eyes closed (n=16)*** |  |  |  |  |  |
| 30s task completed, n | 9 | 9 | 9 | 9 | 1 |
| TIME, s | 26.7 ± 7.0 | 25.57 ± 9.3 | 26.5 ± 12.5 | 28.5 ± 3.9 | **0.046** |
| AREA, mm<sup>2</sup> | 4.37 ± 3.63 | 5.57 ± 4.31 | 11.07 ± 15.29 | 4.35 ± 3.01 | 0.085 |
| VELOCITY |  |  |  |  |  |
| MV, mm/s | 139.06 ± 23.53 | 149.12 ± 38.57 | 192.49 ± 98.08 | 143.03 ± 40.02 | 0.083 |
| MV-AP, mm/s | 16.76 ± 5.28 | 17.72 ± 10.04 | 22.38 ± 12.96 | 17.26 ± 7.34 | 0.167 |
| MV-ML, mm/s | 106.51 ± 61.66 | 72.27 ± 63.99 | 85.49 ± 86.10 | 94.17 ± 75.60 | **0.040** |
| ACCELERATION |  |  |  |  |  |
| ACC, mm/s<sup>2</sup> | 22.23 ± 7.89 | 25.28 ± 10.88 | 30.68 ± 19.53 | 22.19 ± 8.42 | 0.065 |
| ACC-AP, mm/s<sup>2</sup> | 51.73 ± 32.22 | 64.44 ± 38.06 | 57.27 ± 33.94 | 68.90 ± 69.31 | 0.956 |
| ACC-ML, mm/s<sup>2</sup> | 0.98 ± 0.75 | 1.31 ± 1.71 | 1.03 ± 0.57 | 1.70 ± 1.00 | 0.590 |
| JERK |  |  |  |  |  |
| JERK, mm/s<sup>3</sup> | 3.98 ± 2.47 | 3.87 ± 2.15 | 7.29 ± 4.61 | 3.51 ± 0.90 | **0.021** |
| JERK-AP, mm/s<sup>3</sup> | 1.03 ± 0.74 | 2.01 ± 3.49 | 1.74 ± 2.52 | 0.92 ± 1.25 | 0.164 |
| JERK-ML, mm/s<sup>3</sup> | 14.08 ± 7.14 | 17.28 ± 6.86 | 20.56 ± 15.27 | 13.63 ± 5.18 | **0.042** |
| FREQUENCY |  |  |  |  |  |
| MF, Hz | 1.26 ± 0.33 | 1.23 ± 0.37 | 137 ± 0.35 | 1.32 ± 0.33 | 0.845 |

Clinical and functional mobility parameters, and static balance results in tandem and semitandem stances with eyes closed, before and after sham vs real cerebellar rTMS intervention performed in 20 patients with PSP.
**Abbreviations:** ACC, acceleration; AP, anterior-posterior; MF, mean frequency; ML, medio-lateral; mm, millimetre; MV, mean velocity; RMS, Root mean square; s, seconds.

Results from the stance tasks with eyes open showed similar results, although less pronounced. Dynamic mobility assessment did not show differences in performance for real vs sham trial (Supplementary Table 1).

## DISCUSSION

Despite the great advances in the field, postural instability and falls are still important unmet therapeutic targets in PSP [28] and in movement disorders as well [22]. The present trial, using a double-blind sham-controlled crossover design, suggest a beneficial effect of cerebellar rTMS stimulation on measures of postural instability in PSP patients. Our results fit with the observation of alterations of the cerebellum, associated with PSP [8], and with preliminary evidence coming from pilot studies that rTMS may improve functional connectivity between the cerebellum and the motor cortex [9] and that it could improve mobility parameters in PSP [10].

The first published rTMS open-label trial in PSP using cerebellar theta burst stimulation in 10 patients showed an improvement of functional connectivity between cerebellum and motor cortex assessed by neurophysiological measures (i.e. CBI) and functional MRI [9]. However, the trial could not exclude a placebo effect due to the open-label design. Moreover, the authors could not demonstrate a relevant clinical effect. This aspect was recently addressed by a sham-controlled rTMS case study performed for ten days in two PSP patients, showing an improvement in CBI and posturography in the real intervention, although not significant due probably due to an unexpected placebo effect in one patient [10].

The present trial adds novel insights into this concept on multiple levels. First, it provides information about a reasonably large cohort and uses a high quality design. Second, it considered several assessment strategies, including novel mobile health technology, to assess even subtle but potentially clinically relevant parameters. In fact, a single theta burst stimulation showed a surprisingly clear effect on accelerometer-derived measures of static balance, a feature that is regularly affected in PSP and leads to severe impairment in daily activities and quality of life [23,24]. The real intervention showed an effect particularly in the medio-lateral direction of static balance (see, e.g., Table 1 and Supplementary Table 1). This is of interest, as MacKinnon and colleagues [25] and Mitchell and colleagues [26] found that medio-lateral parameters of static balance reflect predominantly axial (e.g., hip and upper thigh) compensatory movements, and antero-posterior parameters predominantly distal (e.g., ankle) compensatory movements. This aspect could be helpful for the design of specific training programs, especially if our results could be confirmed in future studies and an ecological effect (e.g., reduced falls rate) could be associated with rTMS. In our assessments, PSP patients were able to extend the time they could perform the tasks after real stimulation, making us optimistic for the clinical translation of rTMS stimulation. Third, the improvement in static balance, as observed with mobile health technology, was not reflected by any of the clinical and mobility performance test included in the assessment battery.

This result highlights the need for inclusion of such technology in these types of trials, as conventional assessment methods may be too roughly scaled to detect relatively subtle changes [27, 28]. Fourth, the effects of rTMS on static balance parameters were particularly evident in the more challenging tasks. This aspect allows different interpretations and conclusions. It argues for the usefulness of challenging paradigms to be included in assessment panels of clinical trials [29].

Another interpretation could be that PSP patients with relatively preserved static balance performance (and thus potentially lower cerebellar pathology) present with higher functional reserve and thus could show a more effective response to even short stimulations compared to more advanced patients. Indeed, the cohort we selected was in a relatively early disease stage compared to recently published pharmacological trials [30]. The response of a single session rTMS could also identify those patients who might benefit most of longer rTMS trials to be performed in the future. Such trials should include more complex measures (including unsupervised assessments [31]).

In conclusion, this is the first study showing a relevant effect of a short cerebellar rTMS intervention on static balance in PSP patients, supporting the rationale for longer stimulation protocols (e.g., NCT04237948).

## Data Availability

The data will be available for researchers or readers upon reasonable request

## ACKNOWLEDGMENT

The authors thank the study participants and the physiotherapists involved in the study. Without their invaluable contribution, this study would not have been possible.

## AUTHORS’ CONTRIBUTION

1. Research project: A. Conception, B. Organization, C. Execution;
2. Statistical Analysis: A. Design, B. Execution, C. Review and Critique;
3. Manuscript: A. Writing of the first draft, B. Review and Critique

Andrea Pilotto: 1A, 1B, 1C, 2A, 2B, 2C, 3A, 3B.

Maria Cristina Rizzetti: 1C, 2A, 2C, 3A, 3B.

Alberto Lombardi: 1C, 2B, 2C, 3A, 3B.

Clint Hansen: 2B, 2C, 3A, 3B.

Michele Biggi: 1C, 2B.

Giacomo Verzeroli: 1C, 2B.

Antonella Martinelli: 3A, 3B.

Robbin Romijnders: 2B, 2C, 3A, 3B.

Barbara Borroni: 1A, 1B, 2 A, 3A, 3B.

Walter Maetzler: 1A, 1B, 2A, 2B, 2C, 3A, 3B.

Alessandro Padovani: 1A, 1B, 2A, 2B, 2C, 3A, 3B.

---

**Figure 1.**
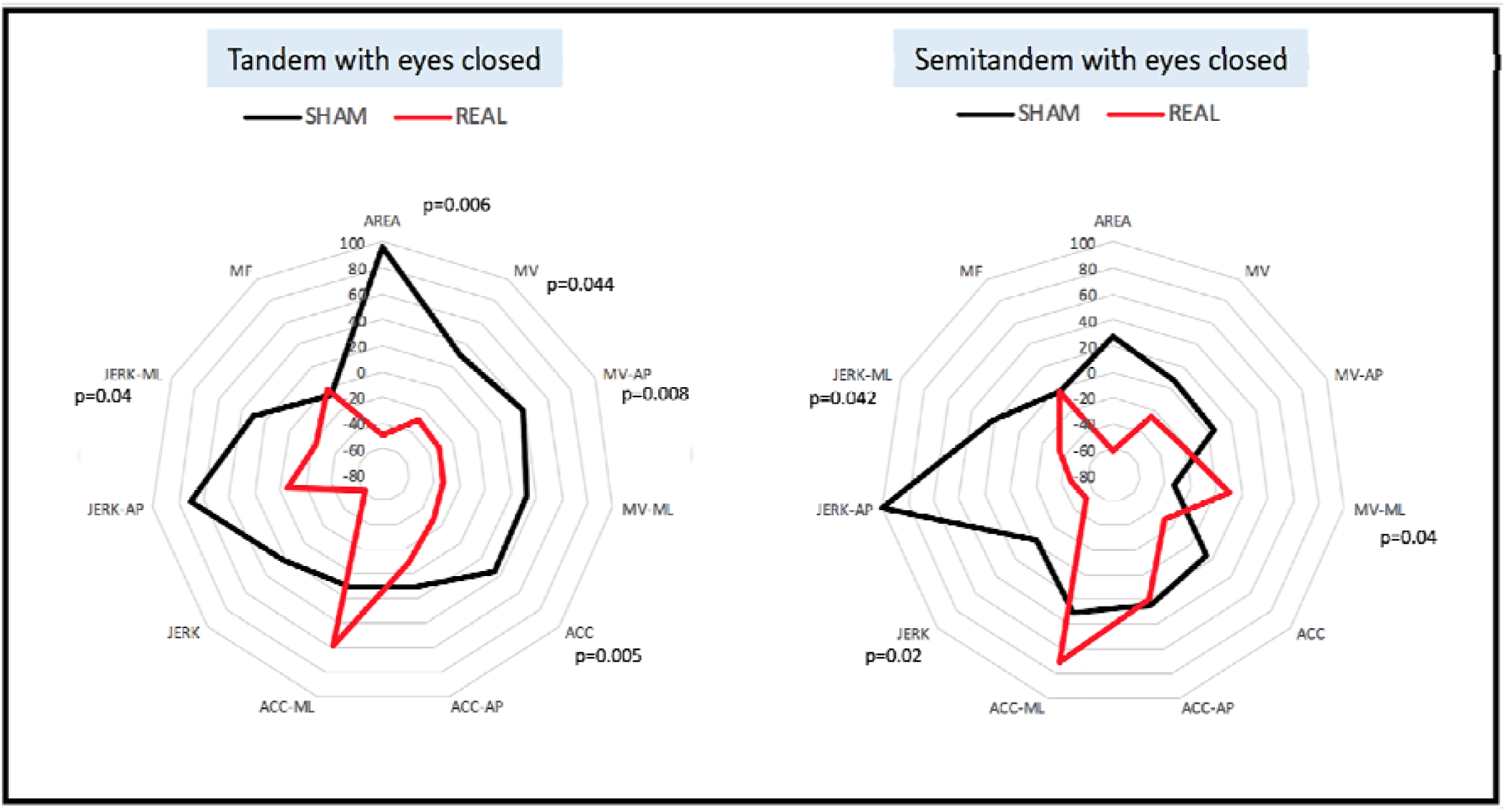
Spider graphs showing the changes (in percentage) of static balance parameters from tandem stance and semitandem stance with eyes closed in patients with PSP, compared to values obtained before the sham versus real intervention. Significant differences are presented with p-values. **Abbreviations:** ACC, acceleration; AP, anterior-posterior; MF, mean frequency; ML, medio-lateral; MV, mean velocity.

