## supplementary material for "Cerebellar rTMS in PSP: a double blind sham-controlled study using mobile health technology"

**Static Balance Assessment**

All patients were supervised in reaching the starting position and arms flagged in self-chosen comfortable position. Each task started with a sound emitted by the tablet, which was only started after adequate instruction and having the OK of the participant that the task can be started. If the participant had to perform a compensatory step during the task, the task was stopped and the time between the sound and the compensatory step was noted. If the participant was not able to maintain the position and did a compensatory step before the task could be started, the task was considered as not being performed due to inability of the participant.

**Postprocessing**

To ensure that only data obtained from the effective static balance phase was included in the analysis, all datasets were 1) counterchecked with the time noted down by the investigator for the specific task and 2) evaluated by visual inspection (CH) to make sure that no step signal as well as no large amplitudes that could not be explained by body sway was included in the final dataset.

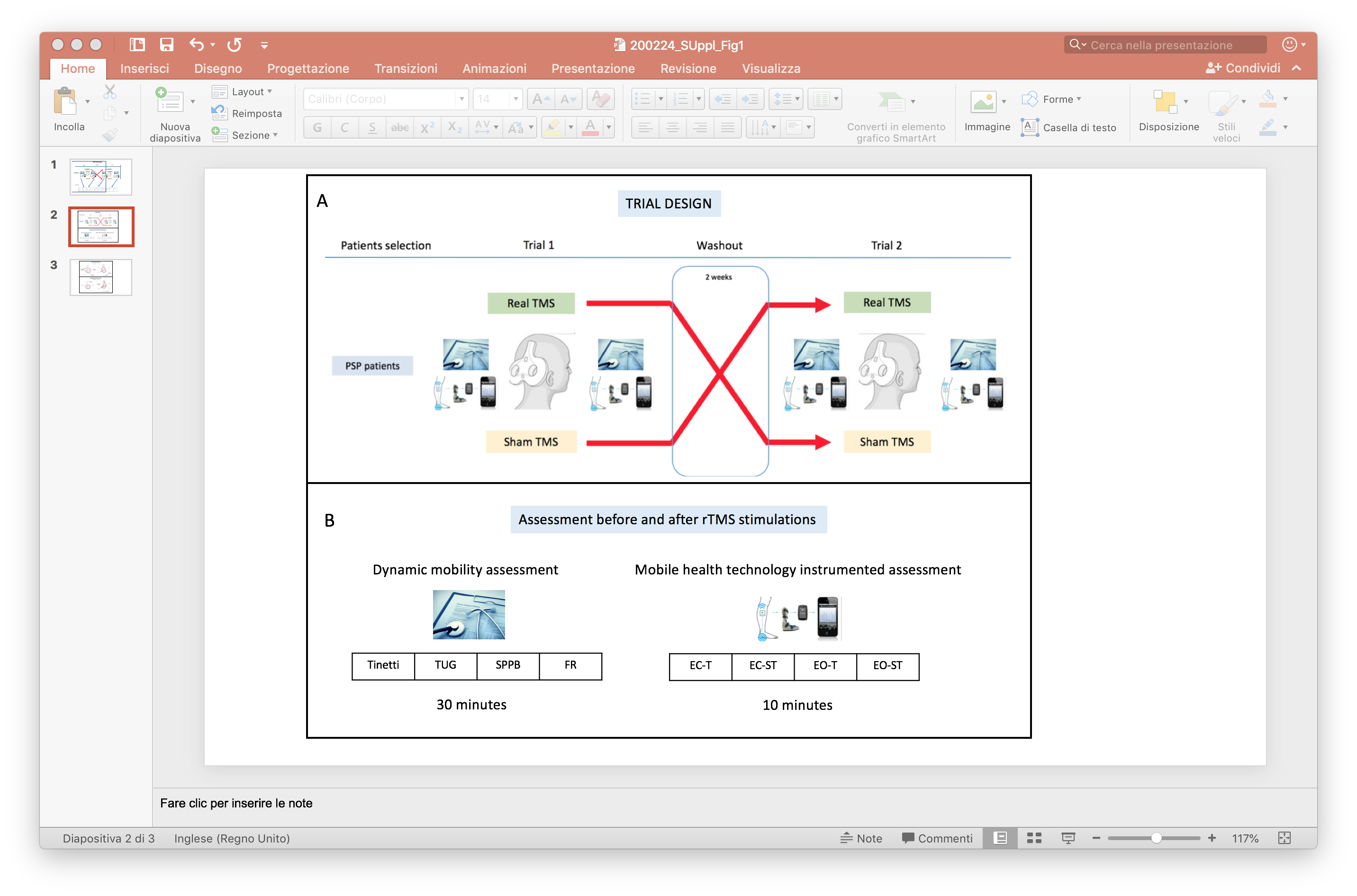

**Supplementary Figure 1 Study design and assessment**

**A** Crossover design of the rTMS study. After recruitment, 10 PSP patients underwent a real rTMS trial, and 10 were sham-treated. After a wash out period of at least two weeks, the second intervention was performed using a crossover design. **B** Assessments were performed before and after each rTMS session (real and sham), and included clinical scales and mobility assessment. Static balance tests were instrumented with mobile health technology (accelerometer device at the lower back, Rehagait®, Hasomed, Germany). EC-ST: semitandem static balance test performed with eyes closed; EC-T, tandem static balance test performed with eyes closed; EO-ST: semitandem static balance test performed with eyes open; EO-T: tandem static balance test performed with eyes open; FR, functional reach test; SPPB, Short Physical Performance Battery; TUG, Timed Up and Go test.

**Supplementary table 1** Results of static balance test in tandem and semitandem position and eyes open.

| **Variable** | **Pre-SHAM** | **Post-SHAM** | **Pre-REAL** | **Post-REAL** | **p** |
| --- | --- | --- | --- | --- | --- |
| ***Tandem stance eyes open (n=20)*** | | | | | |
| 30s task completed, n | 13 | 13 | 13 | 13 | 0.8 |
| TIME, s | 23.1 ± 11.7 | 23.8 ± 10.3 | 23.1 ± 11.7 | 25.3 ± 8.5 | 0.353 |
| AREA, mm^2^ | 6.12 ± 8.62 | 6.55 ± 8.56 | 11.58 ± 16.94 | 7.14 ± 10.50 | 0.423 |
| VELOCITY |  |  |  |  |  |
| MV, mm/s | 147.72 ± 28.66 | 164.68 ± 64.35 | 178.40 ± 63.23 | 147.57 ± 45.65 | **0.050** |
| MV-AP, mm/s | 16.93 ± 10.37 | 18.54 ± 10.76 | 21.83 ± 11.96 | 20.81 ± 21.68 | 0.728 |
| MV-ML, mm/s | 80.31 ± 52.98 | 93.57 ± 83.26 | 111.31 ± 67.16 | 79.43 ± 63.33 | 0.225 |
| ACCELERATION |  |  |  |  |  |
| ACC, mm/s^2^ | 23.93 ± 12.62 | 24.33 ± 13.83 | 32.23 ± 22.34 | 26.62 ± 22.34 | 0.527 |
| ACC-AP, mm/s^2^ | 71.09 ± 37.19 | 42.13 ± 20.37 | 73.64 ± 45.80 | 65.19 ± 57.77 | 0.273 |
| ACC-ML, mm/s^2^ | 1.13 ± 1.14 | 0.83 ± 0.92 | 0.94 ± 1.00 | 1.05 ± 0.73 | 0.358 |
| JERK |  |  |  |  |  |
| JERK, mm/s^3^ | 5.14 ± 4.37 | 6.67 ± 8.05 | 8.84 ± 8.39 | 5.40 ± 5.27 | 0.094 |
| JERK-AP, mm/s^3^ | 2.01 ± 1.56 | 1.35 ± 0.66 | 1.45 ± 1.27 | 1.07 ± 0.82 | 0.660 |
| JERK-ML, mm/s^3^ | 16.62 ± 8.34 | 15.37 ± 9.45 | 23.04 ± 19.76 | 15.40 ± 9.56 | 0.315 |
| FREQUENCY |  |  |  |  |  |
| MF, Hz | 1.36 ± 0.36 | 1.46 ± 0.47 | 1.34 ± 0.45 | 1.41 ± 0.53 | 0.813 |
| ***Semitandem stance eyes open (n=20)*** | | | | | |
| 30s task completed, n | 17 | 17 | 17 | 17 | 0.8 |
| TIME, s | 29.62 ± 1.49 | 28.43 ± 3.44 | 23.1 ± 11.7 | 29.56 ± 1.75 | **0.050** |
| AREA, mm^2^ | 3.63 ± 3.03 | 5.16 ± 8.64 | 8.48 ± 15.62 | 3.53 ± 1.88 | 0.156 |
| VELOCITY |  |  |  |  |  |
| MV, mm/s | 127.48 ± 15.52 | 142.17 ± 40.13 | 145.72 ± 50.57 | 132.71 ± 28.05 | 0.135 |
| MV-AP, mm/s | 15.01 ± 5.12 | 14.47 ± 6.51 | 21.40 ± 26.96 | 14.84 ± 4.03 | 0.385 |
| MV-ML, mm/s | 99.77 ± 60.45 | 66.06 ± 41.70 | 121.79 ± 184.79 | 67.25 ± 23.83 | 0.677 |
| ACCELERATION |  |  |  |  |  |
| ACC, mm/s^2^ | 19.78 ± 6.94 | 22.20 ± 14.00 | 27.20 ± 28.16 | 19.82 ± 5.55 | 0.203 |
| ACC-AP, mm/s^2^ | 60.20 ± 43.89 | 74.13 ± 70.48 | 48.25 ± 35.51 | 74.18 ± 53.35 | 0.588 |
| ACC-ML, mm/s^2^ | 0.93 ± 0.83 | 1.79 ± 0.81 | 1.39 ± 1.11 | 0.73 ± 0.78 | **0.002** |
| JERK |  |  |  |  |  |
| JERK, mm/s^3^ | 3.28 ± 1.30 | 5.57 ± 7.07 | 9.94 ± 18.53 | 3.45 ± 1.97 | 0.104 |
| JERK-AP, mm/s^3^ | 1.03 ± 0.53 | 1.32 ± 1.19 | 1.17 ± 1.02 | 0.98 ± 0.68 | 0.247 |
| JERK-ML, mm/s^3^ | 12.51 ± 5.68 | 16.32 ± 13.16 | 15.61 ± 10.66 | 12.86 ± 4.71 | 0.112 |
| FREQUENCY |  |  |  |  |  |
| MF, Hz | 1.29 ± 0.36 | 1.39 ± 0.44 | 1.361 ± 0.44 | 1.31 ± 0.42 | 0.178 |
| **Clinical assessment and motor tasks** | | | | | |
| Tinetti, total score | 16.9 ± 4.5 | 17.0 ± 4.2 | 16.8 ± 4.5 | 17.5 ± 4.3 | 0.071 |
| TUG, seconds | 20.8 ± 10.5 | 19.9 ± 9.57 | 22.3 ± 9.57 | 21.6 ± 7.41 | 0.882 |
| SPPB, total score | 5.90 ± 2.50 | 5.95 ± 2.48 | 5.85 v 2.41 | 5.90 ± 2.35 | 0.894 |
| Functional reach, cm | 17.1 ± 5.15 | 18.7 ± 6.52 | 17.0 ± 5.17 | 19.0 ± 5.00 | 0.803 |

**Abbreviations:** ACC, acceleration; AP, anterior-posterior; cm, centimetre; FR, functional reach test; MF, mean frequency; ML, medio-lateral; mm, millimeter; MV, mean velocity; RMS, Root mean square, s, seconds; SPPB, Short Physical Performance Battery; TUG, Timed Up and Go test.
